# Cannabinoids Exposure Levels and its Relation to Anxiety Symptoms and Sleep Disturbances: A Scoping Review Protocol

**DOI:** 10.1101/2022.12.15.22283524

**Authors:** Juan Guillermo Perez-Carreño, Krishna Vaddiparti, Elizabeth Castaneda, Gabriela A. Garcia, Pranav Sai Gupta, Catalina Lopez-Quintero

## Abstract

At least 60% of individuals with anxiety disorders report sleep disturbances. Shared physiological mechanisms might explain their co-occurrence. Scientific literature related to medical cannabis, a promising therapeutical candidate for these conditions, increased about 15 times in the last 10 years. However, assessments of cannabinoid exposure, anxiety, and sleep are inconsistent across studies, and the quality of the evidence is not often evaluated. We developed a Scoping Review to examine the current knowledge on these gaps related to cannabinoid use for anxiety and sleep disturbances.

This protocol provides detailed information on how the scoping review will be conducted. It shows the inclusion criteria for studies on the topic of interest as well as the search strategies for the following databases: PubMed, EMBASE, Cochrane Database of Systematic Reviews, Cochrane Central Register of Controlled Trials, CINAHL, LILACS, and PsycINFO. We present the methodological aspects for screening, data extraction, and data charting. In addition, we proposed to evaluate the quality of the evidence by applying critical appraisal tools according to the study designs.

Adherence to this protocol will allow the research team to effectively and reliably synthesize research evidence on the effect of cannabinoids on anxiety symptoms and sleep disturbances.

## Introduction

At least 60% of individuals with anxiety disorders report sleep disturbances, and these disturbances exacerbate symptom severity, increase the likelihood of depression, suicidal behaviors, and substance use disorders, as well as non-responsiveness to treatment(1,2). Common physiological mechanisms can explain the association between sleep disturbances and anxiety, and therefore therapeutic approaches addressing these shared pathways might help improve treatment response, particularly for individuals with anxiety who are non-responsive to conventional treatments(1–3). Looking at new therapeutic targets, recent studies have focused on endocannabinoid system modulation by exogenous cannabinoids, like Cannabidiol (CBD), a promising candidate for wake-sleep regulation with a good safety profile(3–5). Although the interest in medical cannabis products has tripled over the last 10 years, most evidence is based on observational studies that failed to assess cannabis exposure in detail, uncovering difficulties to measure cannabinoid components among the multiple cannabis products in the market. For instance, recent observational studies have focused on describing the population consuming cannabis in terms of sociodemographic characteristics, use of other substances, and the perceived effect of cannabis on anxiety symptoms and sleep quality(6–8). On the other hand, interventional studies have tested specific medical cannabis dosages on a small number of neurologic conditions, like Parkinson’s disease, epilepsy, pain, and Tourette Syndrome(9–12). To the best of our knowledge, evidence regarding the quantification of cannabinoids exposure and its association with anxiety symptoms and sleep disturbances is scarce. Additionally, there is a need to evaluate the quality of the evidence available.

## Aim

This scoping review intends to describe primary studies conducted worldwide on people using cannabis for recreational or medical purposes, establishing the levels of exposure to cannabinoids, its potential relation to anxiety symptoms and sleep disturbances, as well as the risk of bias of such evidence. Linking our results with such information will give us a broader scope of the needs in this field and could led researchers in the field of medical cannabis to refine research questions and methodologies.

### Eligibility criteria

- People whose anxiety and sleep disturbances symptoms are described. This can be detailed as the study inclusion criteria or as outcomes, as soon as give potential association measurement. Either anxiety and sleep can be stated in terms of isolated symptoms with no particular link with a coexisting disease or as a specific diagnosis from the whole spectrum of these entities (i.e. generalized anxiety disorder, panic disorder, and phobias, for anxiety; dyssomnias or parasomnias for sleep-wake disorders). No restriction to diagnosis instruments or methods will be applied.
- Intervention is related to cannabis use in any form (vaping, edible, cream, elixir, and others) and dosage, as long as the exposure dose is clearly described in terms of quantity or frequency. No restriction for comparators and non-exposure groups will be applied.

### Restrictions to eligibility criteria based on study characteristics

- We will include original studies, either with primary or secondary data analysis.
- Study designs will be limited to randomized control trials, quasi-experimental designs, and observational studies with an analytical approach where the association between the exposure and the outcomes has been assessed.
- Observational-descriptive studies will be excluded if the estimates of the association between cannabinoid exposure and the outcomes (anxiety and sleep disturbances) do not provide sufficient causal evidence.
- No restrictions will apply to the publication date or sample size.

### Restrictions to eligibility criteria based on sources of information

- Publication status will be restricted to fully published papers or in-press papers. Conference abstracts will not be considered because 1) their limited information for risk of bias assessment; 2) requesting missing information from the authors for risk of bias assessment is not usual practice and can lead to misinterpretations and non-replicable approaches.
- No language restriction will be applied.
- Studies subjects will be restricted to humans.

### Search strategy

We used cannabis, marijuana, anxiety, and sleep as terms related to our key concepts. We structured a search strategy based on variations of the terms and other associated terms. For example, for cannabis we used terms related to medical and non-medical cannabis, as well as registered medical cannabis names; for anxiety, we included terms related to the whole spectrum of anxiety-related symptoms and disorders, and terms related to diagnostic tools; finally, for sleep, we included terms related to the sleep-wake cycle, dyssomnias, parasomnias, and instruments to measure sleep disturbances and sleep phases. Expert consultation was carried out to add new terms related to our objective. Published key papers were used to explore more terms associated. We used the database’s thesaurus, where available, and adapted the search strategy to each database and electronic source. PubChem for related terms and free terms were also included. Using Boolean operators, the search strategy was an intersection of the three key concept terms. Two reviewers discussed the search strategy to evaluate its coherence and concordance with our scope. Preliminary and iterative searches were run in PubMed during the whole search strategy development prior to the final terms combinations being adopted. The databases used for retrieving the studies included: PubMed, EMBASE, Cochrane Database of Systematic Reviews, Cochrane Central Register of Controlled Trials, CINAHL, LILACS, and PsycINFO. See complete search strategies in appendix 1. No restrictions based con date, publication format, or language were applied for the search strategies in PubMed, Cochrane Database of Systematic Reviews, Cochrane Central Register of Controlled Trials, CINAHL, LILACS, and PsycINFO. For EMBASE, restrictions were applied to publication types, to retrieve published and in-press articles only.

### Other methods used to identify relevant studies

We consulted experts in the field to identify potential studies conducted or in course related to our key concepts. Included studies in the systematic literature reviews retrieved from our search strategies were checked manually. Review articles related to our key concepts were used to check the reference list, manually. Randomized control trial protocols will be examined for final result publications.

### Study selection

Retrieved documents will be compiled on Zotero to detect and eliminate duplicates. The screening process will be carried out using Rayyan. Four reviewers, independently, will begin reading titles and abstracts from the first 5% of documents as a calibration process, checking they match our inclusion criteria. We pre-specified an agreement threshold of 80% between the reviewers to indicate moderate intersubject correlation. After discussing disagreements among 5 reviewers, we will increase by 5% each calibration round until reaching our correlation threshold. The complete screening process will be carried out by four reviewers, guaranteeing each paper will be read at least by two reviewers. Disagreements will be discussed and resolved among 5 reviewers. Excluded studies will be categorized as not matching the inclusion criteria. A similar calibration and screening process for full documents will be conducted, guaranteeing each paper will be read at least by two reviewers, independently. After checking the inclusion criteria, selected documents will be excluded for the following reasons: Other design than specified by inclusion criteria; other publication types different than specified by inclusion criteria; population not described in terms of anxiety symptoms or sleep-wake disturbance; intervention or exposure different than cannabis for at least 30% of the study population (considering control or comparison group); cannabis, marijuana or cannabinoid dose exposure not stated in terms of frequency and quantity; no association described between cannabis exposure and anxiety symptoms or sleep-wake disturbances. The results for the screening process will be synthesized using the Preferred Reporting Items for Systematic reviews and Meta-Analyses (PRISMA) Flow Diagram.

### Data collection

Two reviewers independently will extract data from selected studies using structured data extraction forms. The information will include general study data (publication year, countries where the study was conducted, funding), the design used (cross-sectional, case-control studies, cohort studies, interventional studies), population characteristics (sample size, health condition of interest, age, sex), methods for evaluating anxiety and sleep disturbance, intervention or exposure of interest (type of cannabinoid, dosage, routes of administration), control or comparator group characteristics, and outcomes (estimated effect size, drug-drug interaction when reported). A synchronic and electronic form will be tested on two studies randomly selected (one observational and one interventional study) and reviewed by at least two reviewers, independently. Changes in the format will be applied after a group discussion. Each reviewer will complete the synchronic form by reading the full document assigned. Authors will be contacted in case of missing information.

### Quality assessment

We acknowledge the risk of bias and internal validity assessment is not a crucial component for scoping reviews. However, we planned overall quality assessment as part of our research question. Two reviewers, independently, will apply critical appraisal tools according to the study design: Cochrane risk-of-bias tool (RoB 2) for randomized trials, Risk Of Bias In Non-Randomized Studies - of Interventions (ROBINS-I) for non-randomized studies, and Joanna Briggs Institute Critical Appraisal tools for observational studies. Results will be discussed in group sessions to solve discrepancies by consensus.

### Data charting

Extracted data will be summarized in tables and figures following our key concept structure. We will present information according to the main population involved in the study, as follows: 1) individuals selected on the basis of having and anxiety-related symptoms <u>and</u> sleep-wake disturbances, 2) individuals selected on the basis of only having anxiety-related symptoms (sleep-wake disturbances were also assessed as an exposure or outcome), 3) individuals selected on the basis of only having sleep-wake disturbances (anxiety-related symptoms were also reported as an exposure or outcome), 4) individuals whose anxiety-related outcome and sleep-week disturbances were not reported as a baseline or inclusion criteria but were reported as secondary exposures or outcomes. Associated quality assessment information will be linked to each study. We will explore tables and figures for each case and will decide to report either the table or figure as they lead to a more comprehensive understanding. We will adhere to the PRISMA extension for scoping reviews(13) to conduct this scoping review and report the final results.

### Review Status

By the time of this report, search strategies were all run on the selected databases, and the screening process by reading titles and abstracts had started. This initial screening helped to refine our inclusion criteria and improve the protocol. The final report will include the role of each reviewer and researcher in the process.

## Data Availability

All data produced in the present study will be available upon reasonable request to the authors.

## Funding

There is no specific funding for this study.

## Competing interests

Reviewers have reported not having any secondary interest that potentially competes with the primary interest of this scoping review.

## Appendix

### Search Strategies

Database: PubMed. Date: October 2^nd^, 2022.

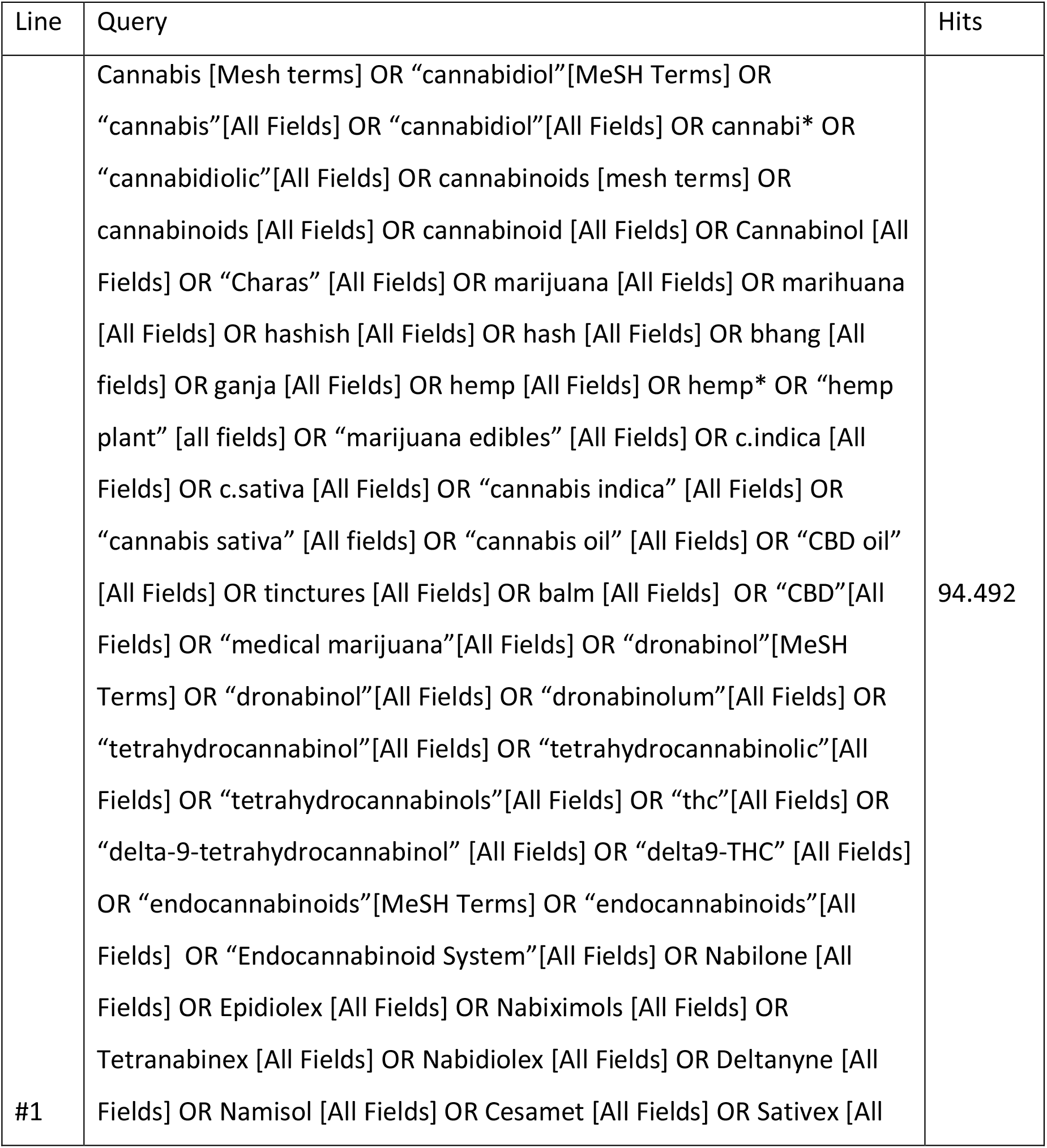
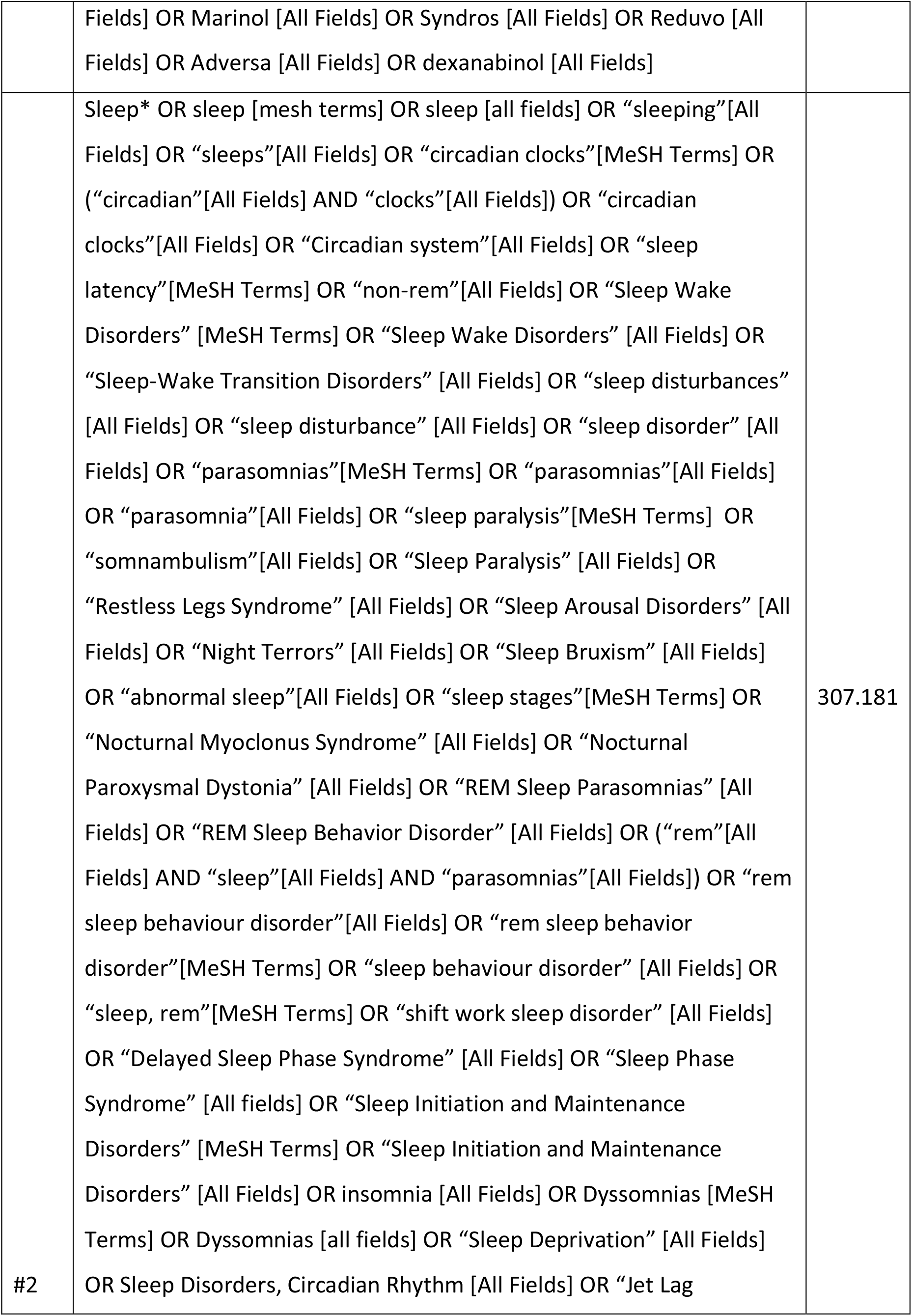
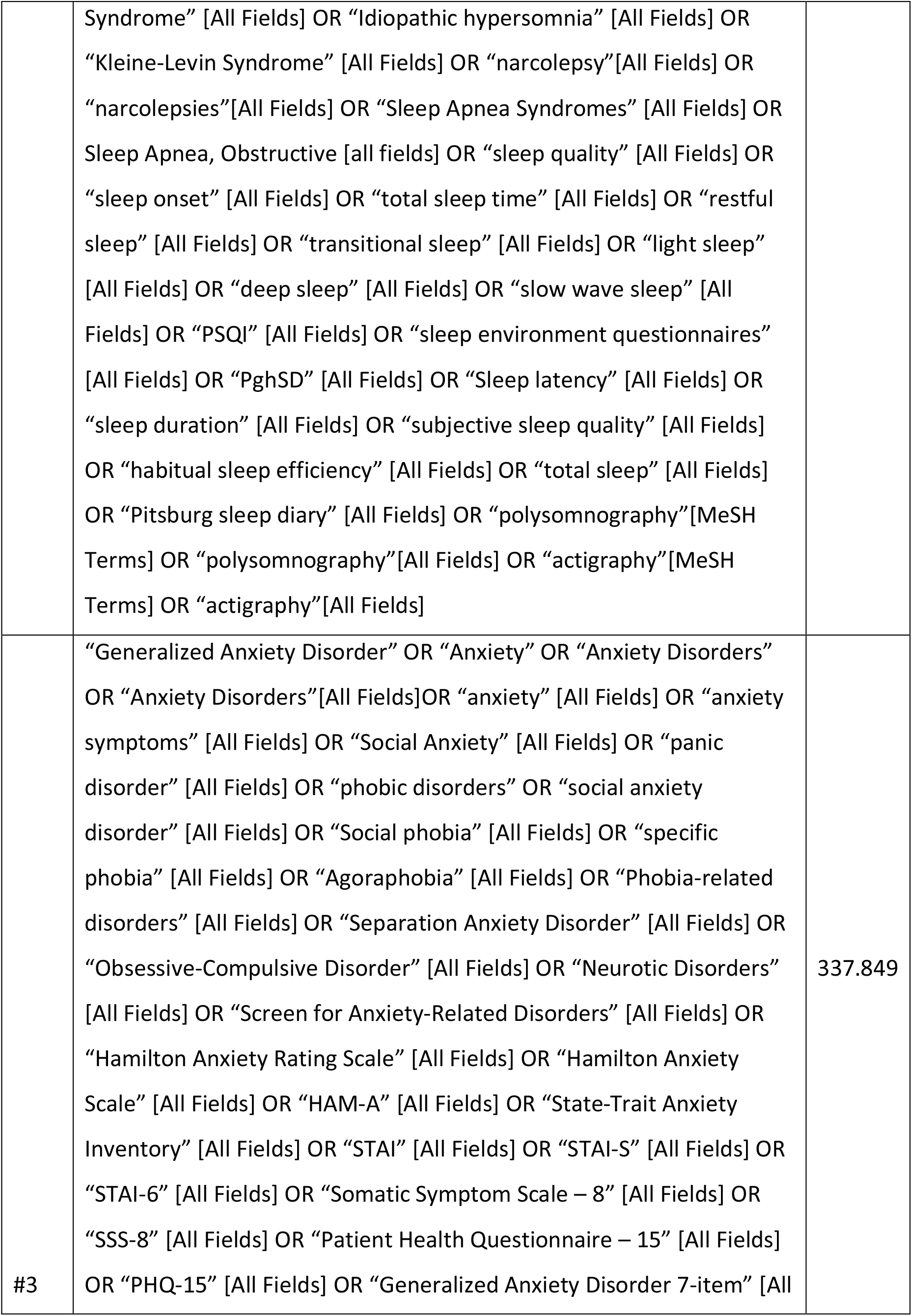
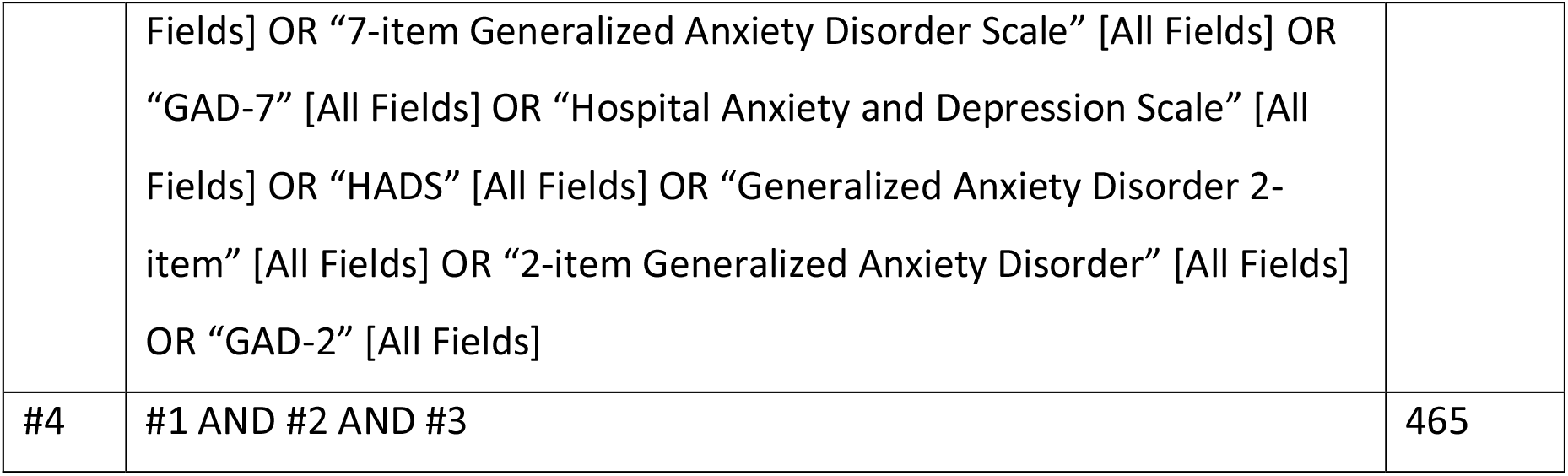

Database: EMBASE. Date: October 2^nd^, 2022.

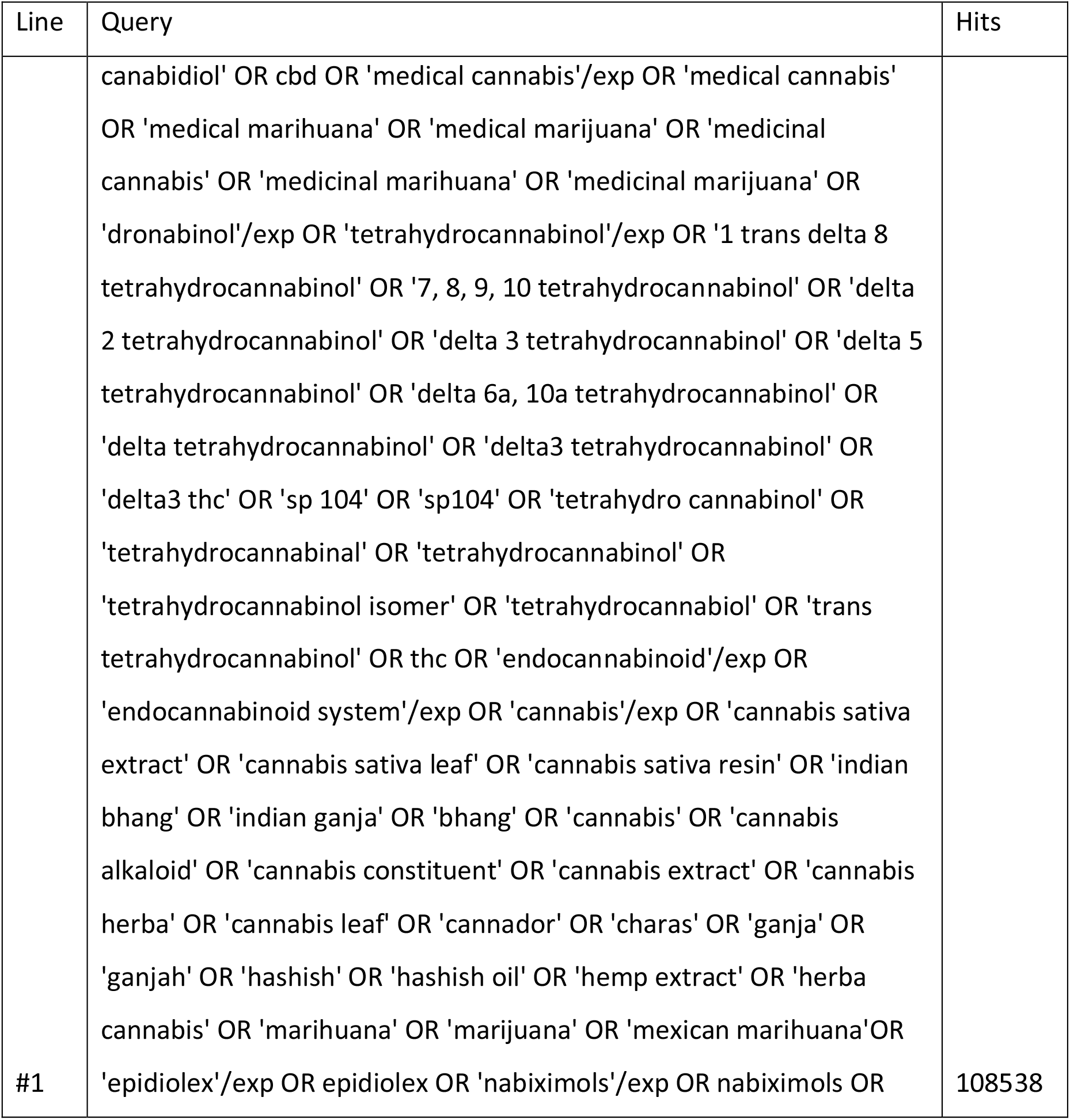
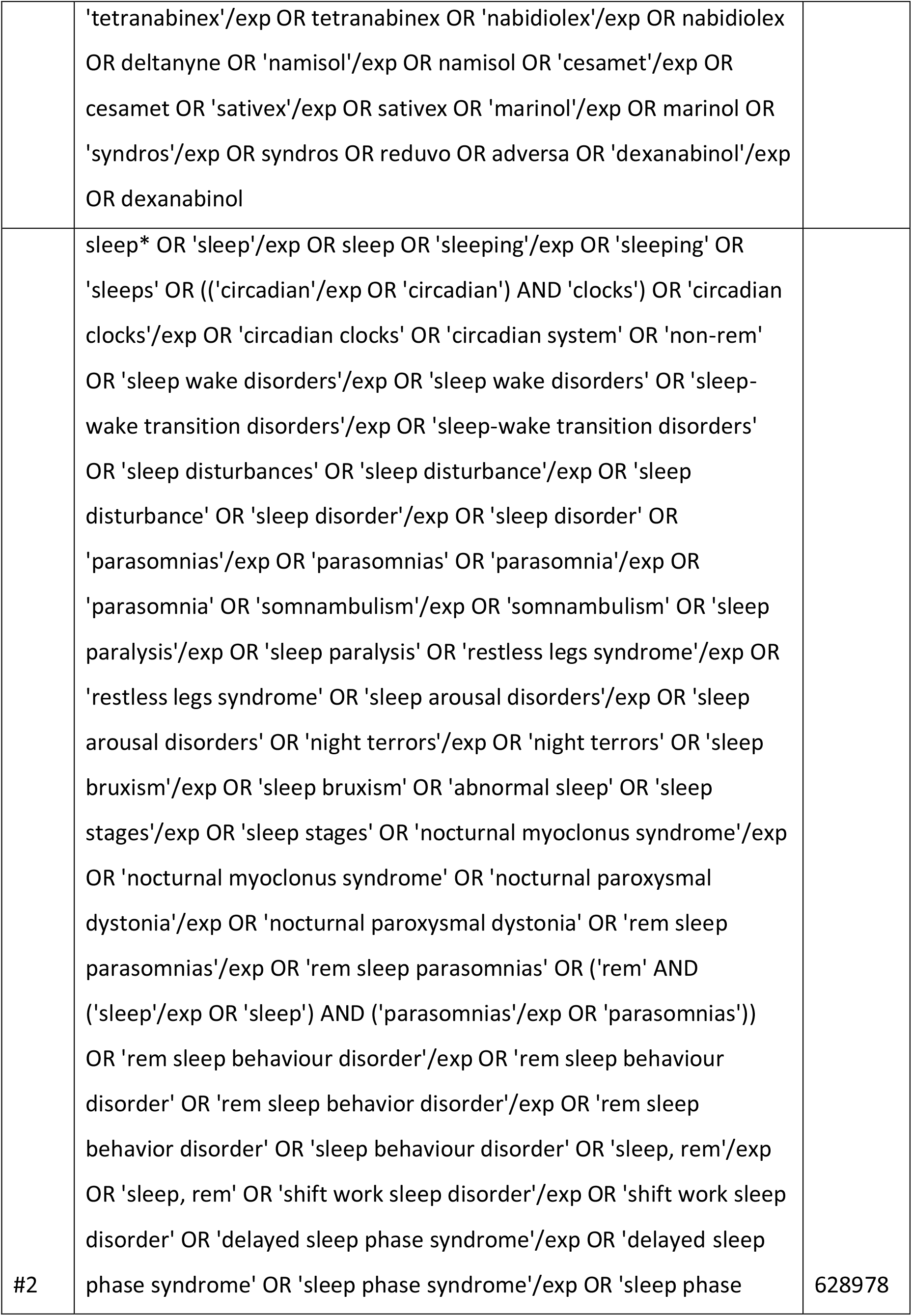
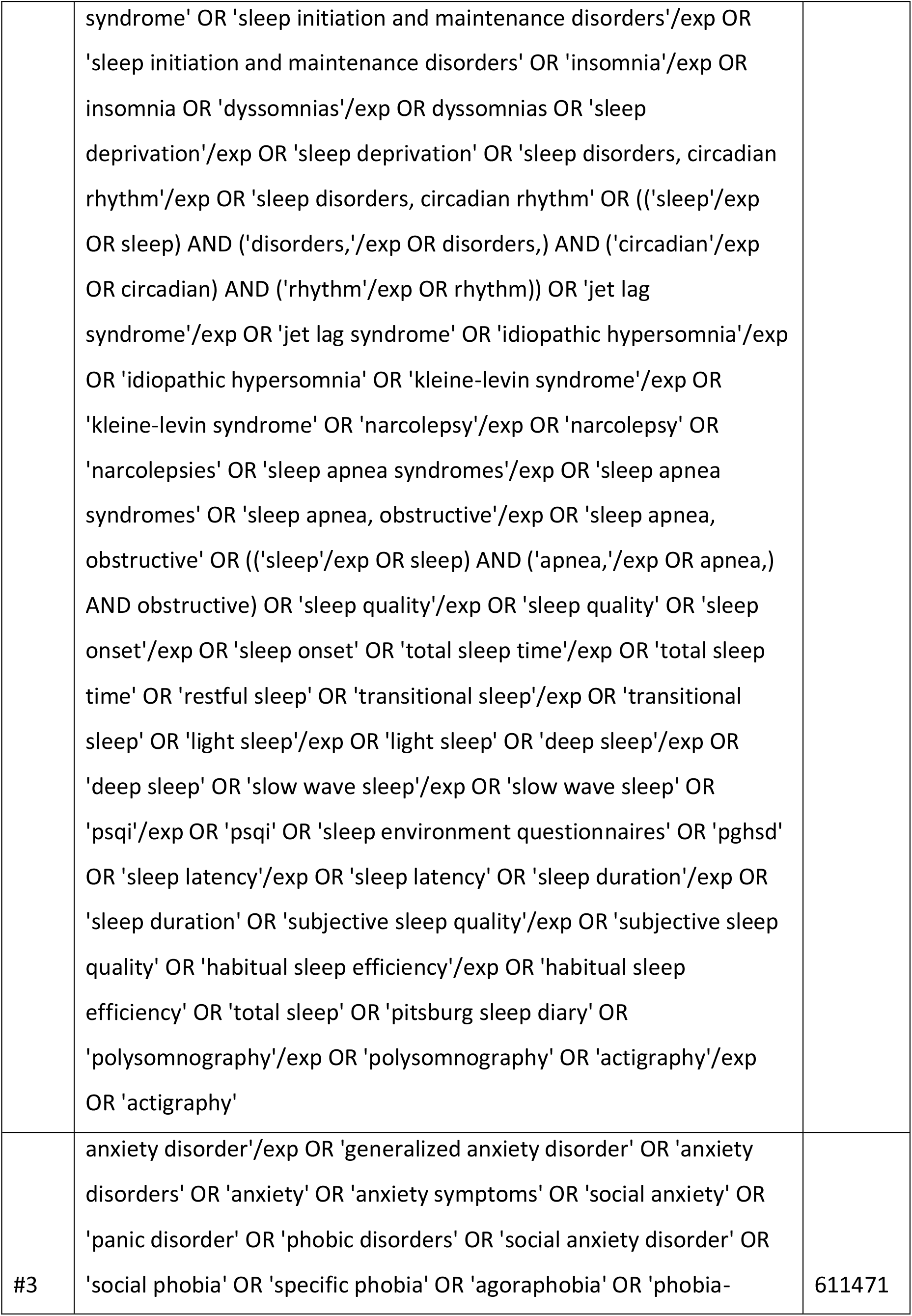
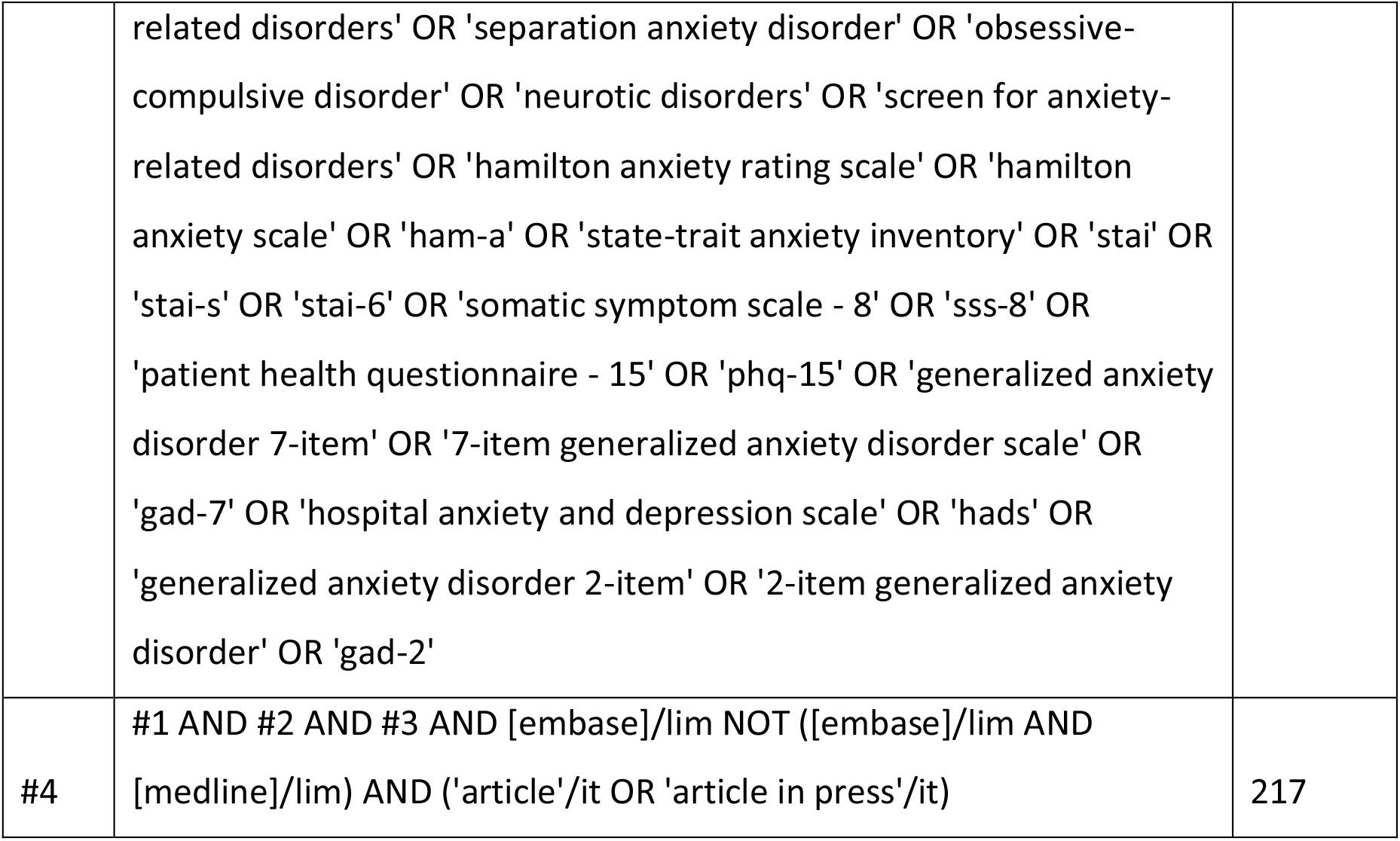

Database: Cochrane Database of Systematic Reviews. Date: October 2^nd^, 2022.

| Line | Query | Hits |
| --- | --- | --- |
| #1 | Cannabis OR "cannabidiol" OR "cannabis" OR "cannabidiol" OR cannabi* OR "cannabidiolic" OR cannabinoids OR cannabinoids OR cannabinoid OR Cannabinol OR "Charas" OR marijuana OR marihuana OR hashish OR hash OR bhang OR ganja OR hemp OR hemp* OR "hemp plant" OR "marijuana edibles" OR c.indica OR c.sativa OR "cannabis indica" OR "cannabis sativa" OR "cannabis oil" OR "CBD oil" OR tinctures OR balm OR "CBD" OR "medical marijuana" OR "dronabinol" OR "dronabinol" OR "dronabinolum" OR "tetrahydrocannabinol" OR "tetrahydrocannabinolic" OR "tetrahydrocannabinols" OR "thc" OR "delta-9-tetrahydrocannabinol" OR "delta9-THC" OR "endocannabinoids" OR "endocannabinoids" OR "Endocannabinoid System" OR Nabilone OR Epidiolex OR Nabiximols OR Tetranabinex OR Nabidiolex OR Deltanyne OR Namisol OR | 4968 |
|  | Cesamet OR Sativex OR Marinol OR Syndros OR Reduvo OR Adversa<br>OR dexanabinol |  |
| #2 | Sleep* OR sleep OR sleep OR "sleeping" OR "sleeps" OR "circadian<br>clocks" OR ("circadian" AND "clocks") OR "circadian clocks" OR<br>"Circadian system" OR "sleep latency" OR "non-rem" OR "Sleep Wake<br>Disorders" OR "Sleep Wake Disorders" OR "Sleep-Wake Transition<br>Disorders" OR "sleep disturbances" OR "sleep disturbance" OR<br>"sleep disorder" OR "parasomnias" OR "parasomnias" OR<br>"parasomnia" OR "sleep paralysis" OR "somnambulism" OR "Sleep<br>Paralysis" OR "Restless Legs Syndrome" OR "Sleep Arousal Disorders"<br>OR "Night Terrors" OR "Sleep Bruxism" OR "abnormal sleep" OR<br>"sleep stages" OR "Nocturnal Myoclonus Syndrome" OR "Nocturnal<br>Paroxysmal Dystonia" OR "REM Sleep Parasomnias" OR "REM Sleep<br>Behavior Disorder" OR ("rem" AND "sleep" AND "parasomnias") OR<br>"rem sleep behaviour disorder" OR "rem sleep behavior disorder" OR<br>"sleep behaviour disorder" OR "sleep, rem" OR "shift work sleep<br>disorder" OR "Delayed Sleep Phase Syndrome" OR "Sleep Phase<br>Syndrome" OR "Sleep Initiation and Maintenance Disorders" OR<br>"Sleep Initiation and Maintenance Disorders" OR insomnia OR<br>Dyssomnias OR Dyssomnias OR "Sleep Deprivation" OR Sleep<br>Disorders, Circadian Rhythm OR "Jet Lag Syndrome" OR "Idiopathic<br>hypersomnia" OR "Kleine-Levin Syndrome" OR "narcolepsy" OR<br>"narcolepsies" OR "Sleep Apnea Syndromes" OR Sleep Apnea,<br>Obstructive OR "sleep quality" OR "sleep onset" OR "total sleep<br>time" OR "restful sleep" OR "transitional sleep" OR "light sleep" OR<br>"deep sleep" OR "slow wave sleep" OR "PSQI" OR "sleep<br>environment questionnaires" OR "PghSD" OR "Sleep latency" OR<br>"sleep duration" OR "subjective sleep quality" OR "habitual sleep<br>efficiency" OR "total sleep" OR "Pittsburg sleep diary" OR | 394 |
|  | "polysomnography" OR "polysomnography" OR "actigraphy" OR "actigraphy" |  |
| #3 | "Generalized Anxiety Disorder" OR "Anxiety" OR "Anxiety Disorders" OR "Anxiety Disorders" OR "anxiety" OR "anxiety symptoms" OR "Social Anxiety" OR "panic disorder" OR "phobic disorders" OR "social anxiety disorder" OR "Social phobia" OR "specific phobia" OR "Agoraphobia" OR "Phobia related disorders" OR "Separation Anxiety Disorder" OR "Obsessive Compulsive Disorder" OR "Neurotic Disorders" OR "Screen for Anxiety Related Disorders" OR "Hamilton Anxiety Rating Scale" OR "Hamilton Anxiety Scale" OR "HAM A" OR "State Trait Anxiety Inventory" OR "STAI" OR "STAI S" OR "STAI 6" OR "Somatic Symptom Scale 8" OR "SSS 8" OR "Patient Health Questionnaire 15" OR "PHQ 15" OR "Generalized Anxiety Disorder 7 item" OR "7 item Generalized Anxiety Disorder Scale" OR "GAD 7" OR "Hospital Anxiety and Depression Scale" OR "HADS" OR "Generalized Anxiety Disorder 2 item" OR "2 item Generalized Anxiety Disorder" OR "GAD 2" | 5063 |
| #4 | #1 AND #2 AND #3 | 74 |

Database: Cochrane Central Register of Controlled Trials. Date: October 2^nd^, 2022.

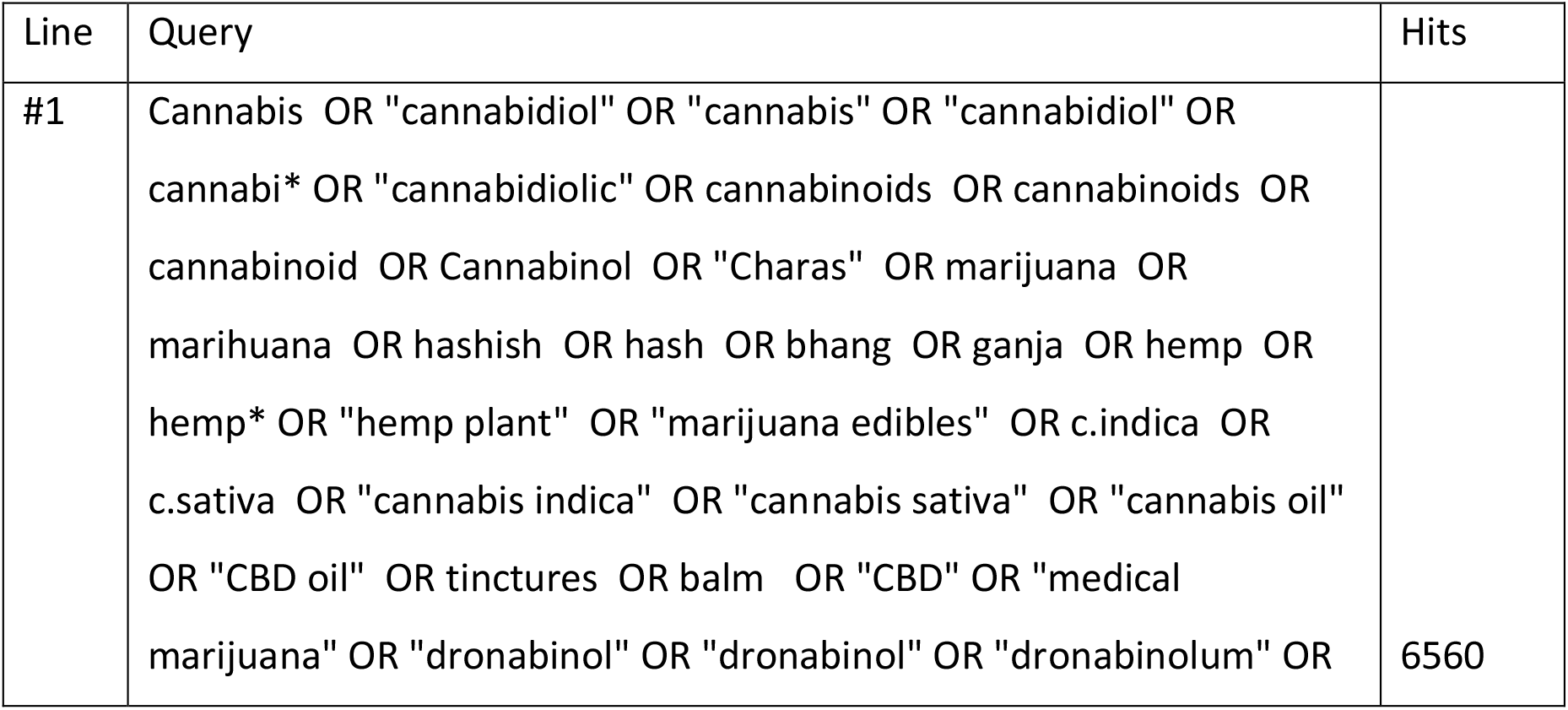
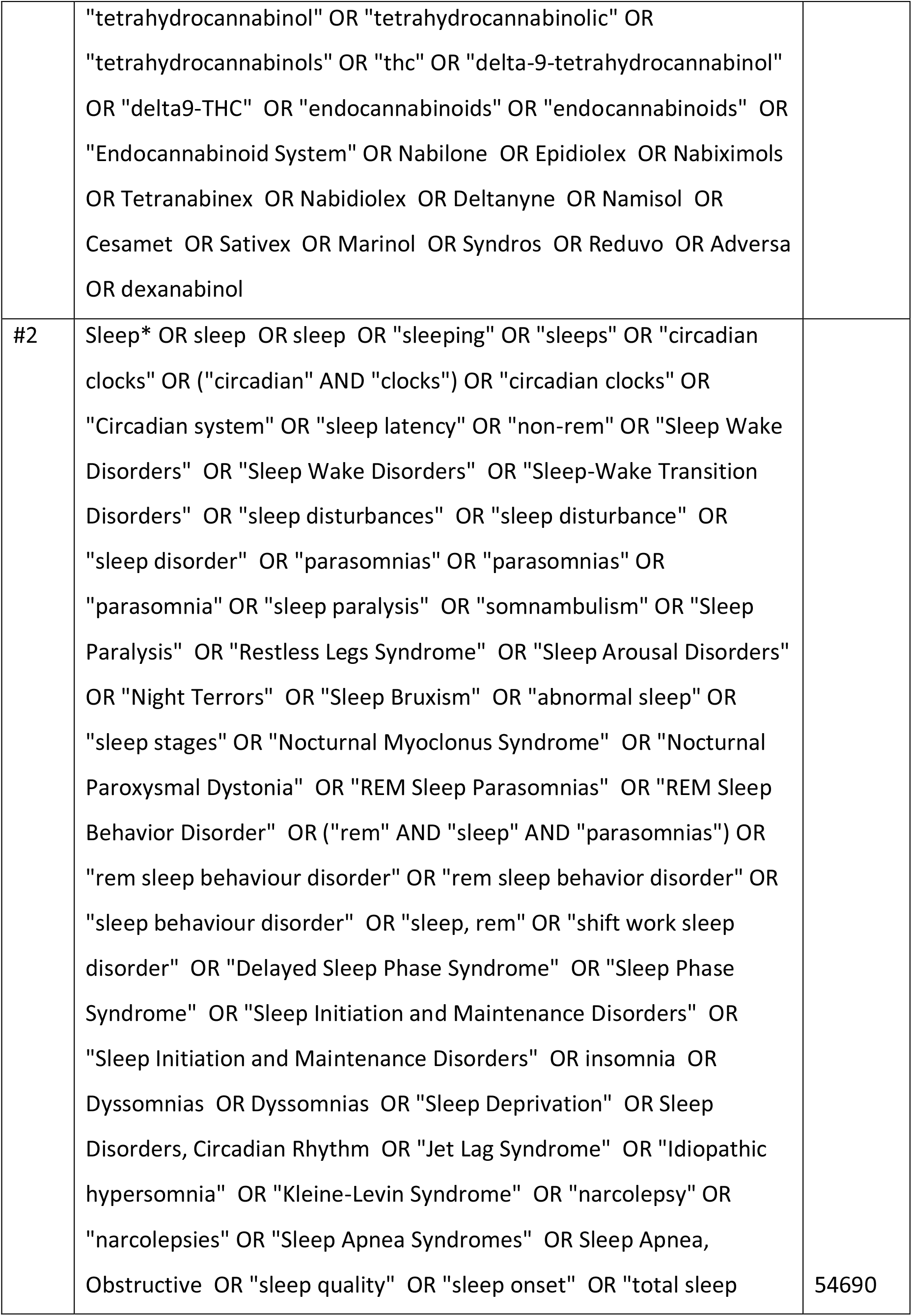
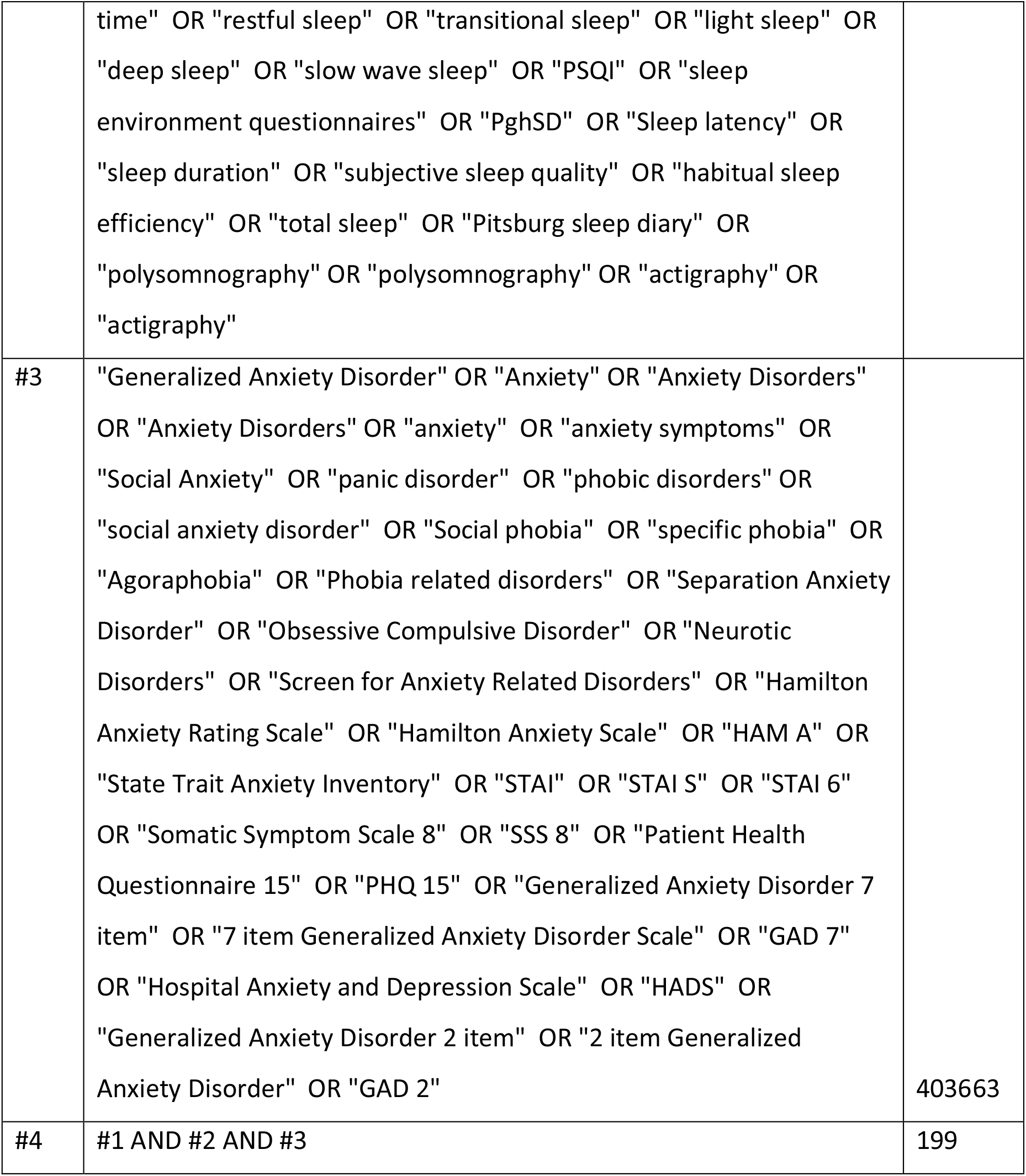

Database: CINAHL. Date: October 2^nd^, 2022. Interface: EBSCOhost Research Databases.

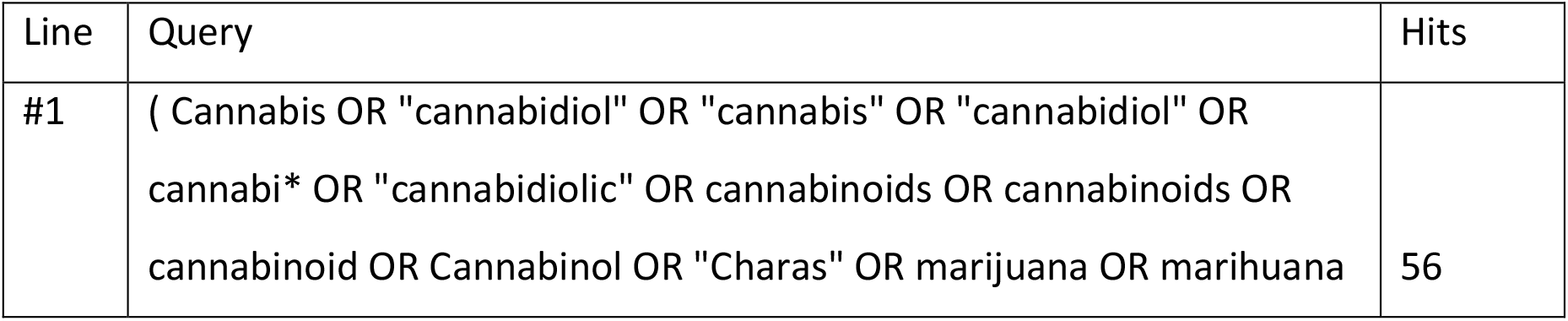
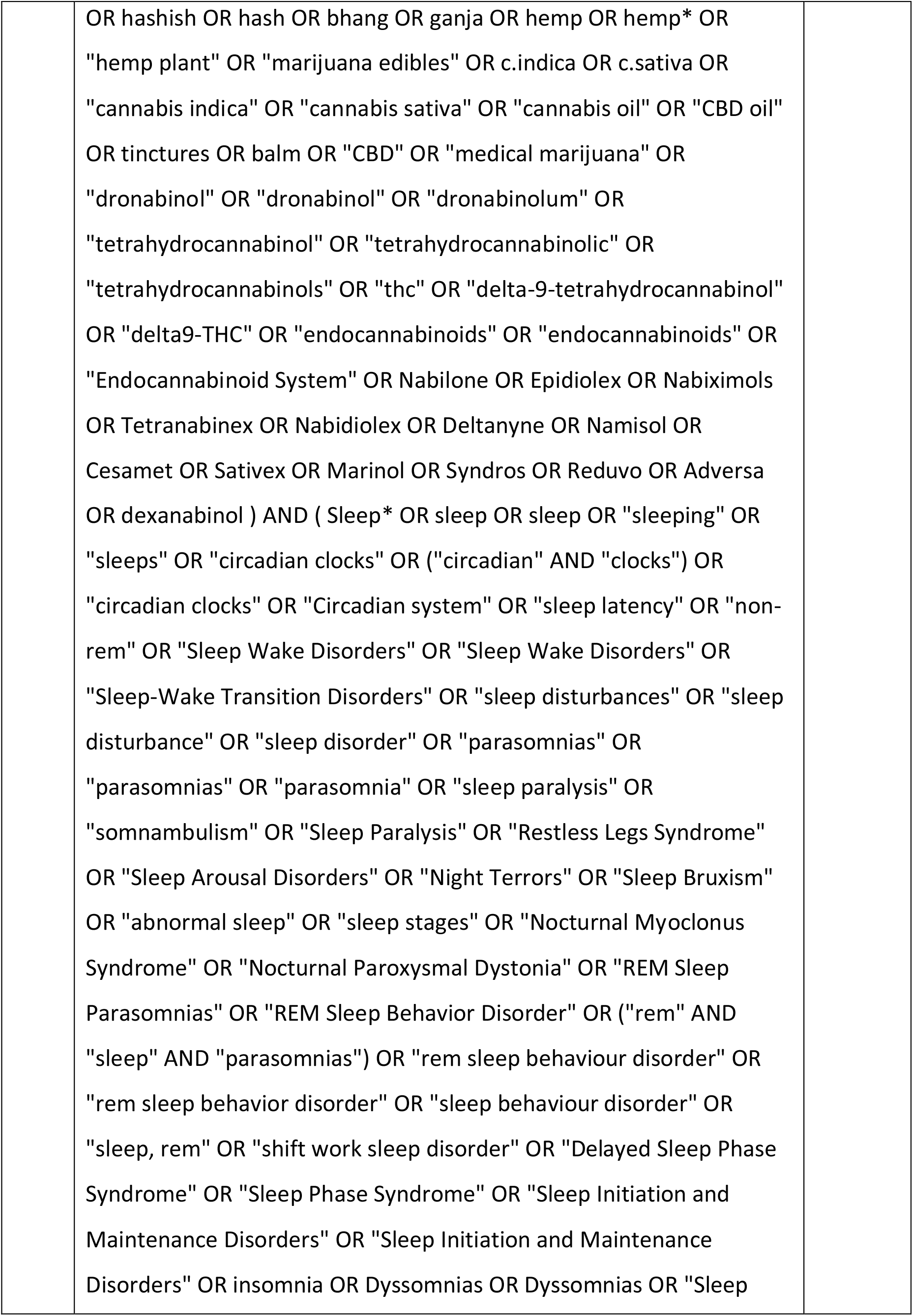
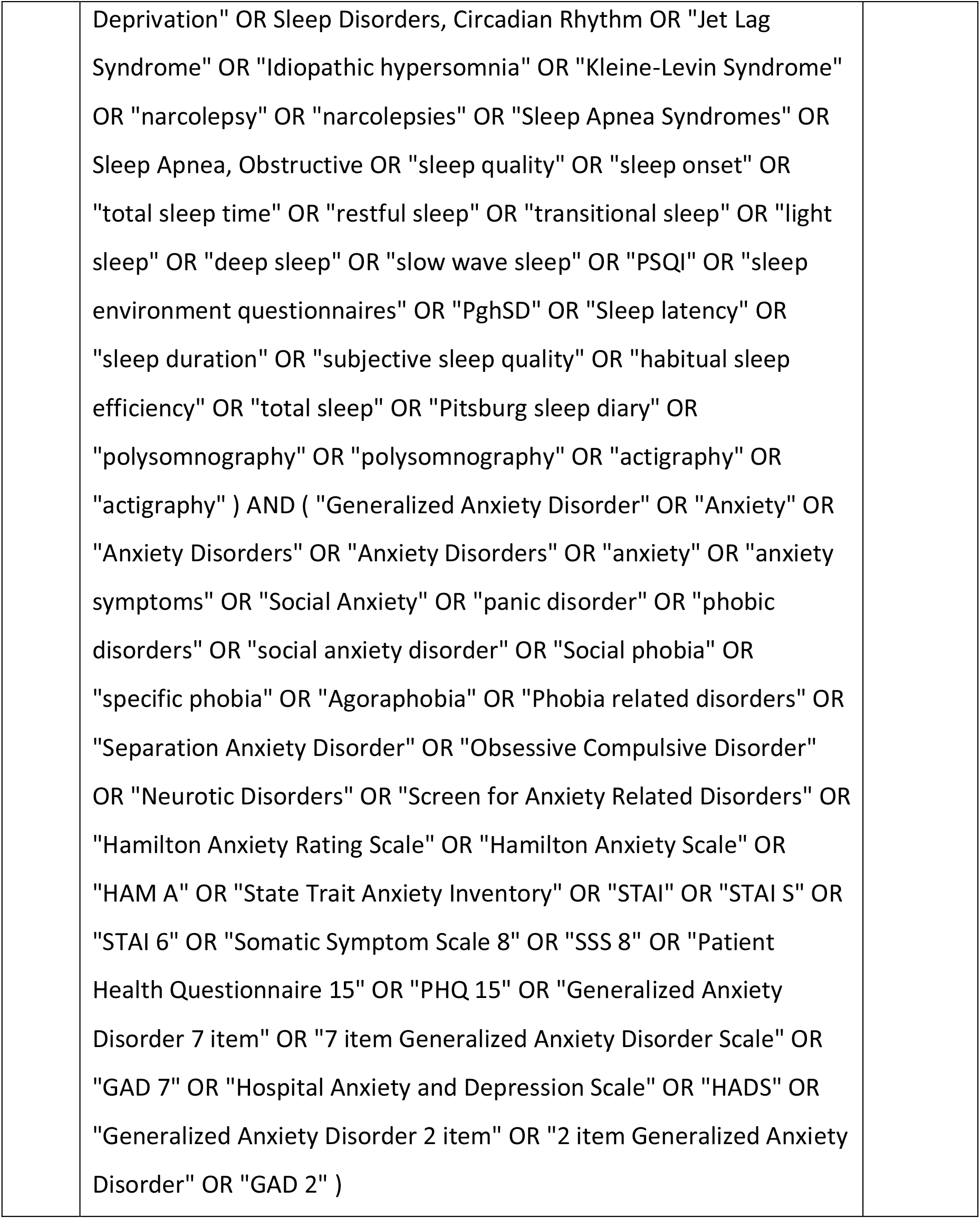

Database: LILACS. Date: October 2^nd^, 2022.

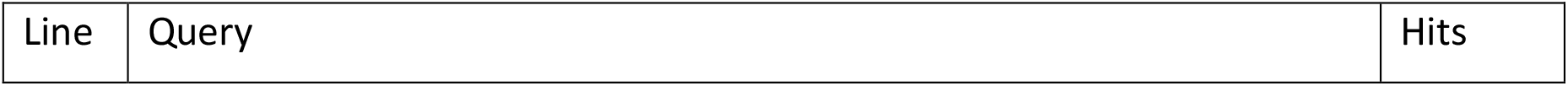
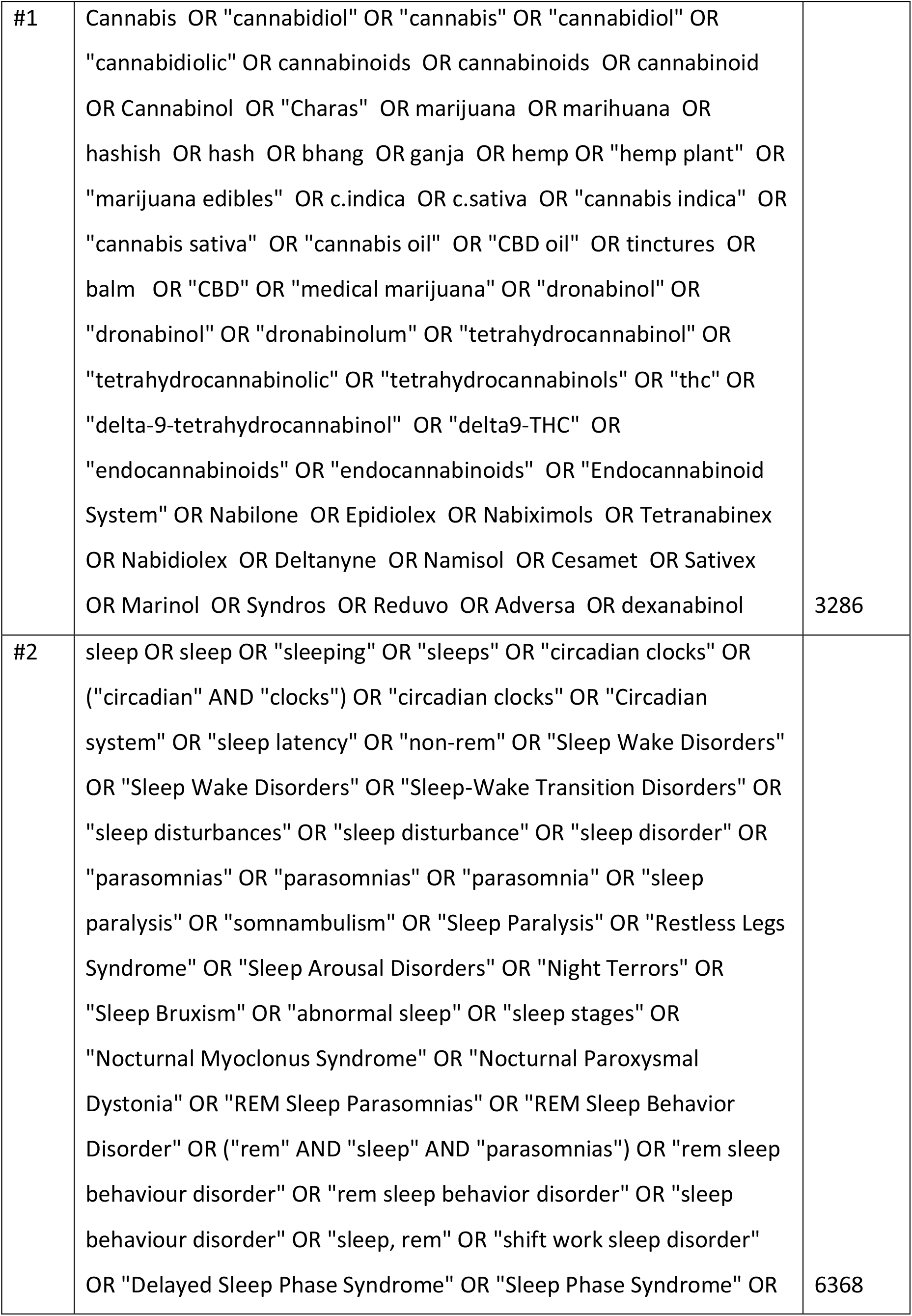
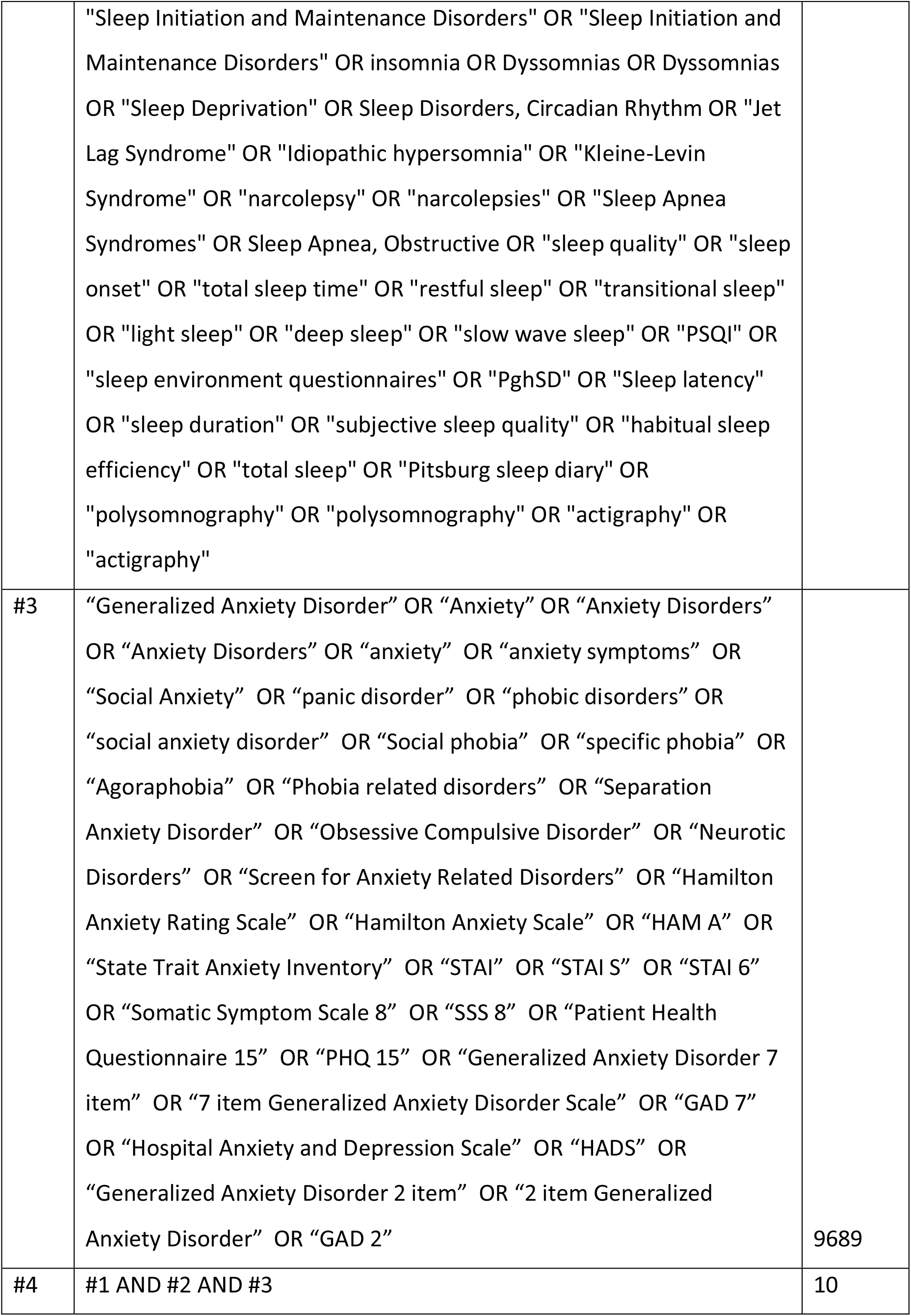

Database: PsycInfo. Date: November 25^th^, 2022. Interface: EBSCOhost Research Databases.

| Line | Query | Hits |
| --- | --- | --- |
| #1 | (Cannabis OR "cannabidiol" OR "cannabis" OR "cannabidiol" OR cannabi* OR "cannabidiolic" OR cannabinoids OR cannabinoids OR cannabinoid OR Cannabinol OR "Charas" OR marijuana OR marihuana OR hashish OR hash OR bhang OR ganja OR hemp OR hemp* OR "hemp plant" OR "marijuana edibles" OR c.indica OR c.sativa OR "cannabis indica" OR "cannabis sativa" OR "cannabis oil" OR "CBD oil" OR tinctures OR balm OR "CBD" OR "medical marijuana" OR "dronabinol" OR "dronabinol" OR "dronabinolum" OR "tetrahydrocannabinol" OR "tetrahydrocannabinolic" OR "tetrahydrocannabinols" OR "thc" OR "delta-9-tetrahydrocannabinol" OR "delta9-THC" OR "endocannabinoids" OR "endocannabinoids" OR "Endocannabinoid System" OR Nabilone OR Epidiolex OR Nabiximols OR Tetranabinex OR Nabidiolex OR Deltanyne OR Namisol OR Cesamet OR Sativex OR Marinol OR Syndros OR Reduvo OR Adversa OR dexanabinol ) AND ( Sleep* OR sleep OR sleep OR "sleeping" OR "sleeps" OR "circadian clocks" OR ("circadian" AND "clocks") OR "circadian clocks" OR "Circadian system" OR "sleep latency" OR "non-rem" OR "Sleep Wake Disorders" OR "Sleep Wake Disorders" OR "Sleep-Wake Transition Disorders" OR "sleep disturbances" OR "sleep disturbance" OR "sleep disorder" OR "parasomnias" OR "parasomnias" OR "parasomnia" OR "sleep paralysis" OR "somnambulism" OR "Sleep Paralysis" OR "Restless Legs Syndrome" OR "Sleep Arousal Disorders" OR "Night Terrors" OR "Sleep Bruxism" OR "abnormal sleep" OR "sleep stages" OR "Nocturnal Myoclonus Syndrome" OR "Nocturnal Paroxysmal Dystonia" OR "REM Sleep | 242 |
|  | Parasomnias" OR "REM Sleep Behavior Disorder" OR ("rem" AND "sleep" AND "parasomnias") OR "rem sleep behaviour disorder" OR "rem sleep behavior disorder" OR "sleep behaviour disorder" OR "sleep, rem" OR "shift work sleep disorder" OR "Delayed Sleep Phase Syndrome" OR "Sleep Phase Syndrome" OR "Sleep Initiation and Maintenance Disorders" OR "Sleep Initiation and Maintenance Disorders" OR insomnia OR Dyssomnias OR Dyssomnias OR "Sleep Deprivation" OR Sleep Disorders, Circadian Rhythm OR "Jet Lag Syndrome" OR "Idiopathic hypersomnia" OR "Kleine-Levin Syndrome" OR "narcolepsy" OR "narcolepsies" OR "Sleep Apnea Syndromes" OR Sleep Apnea, Obstructive OR "sleep quality" OR "sleep onset" OR "total sleep time" OR "restful sleep" OR "transitional sleep" OR "light sleep" OR "deep sleep" OR "slow wave sleep" OR "PSQI" OR "sleep environment questionnaires" OR "PghSD" OR "Sleep latency" OR "sleep duration" OR "subjective sleep quality" OR "habitual sleep efficiency" OR "total sleep" OR "Pittsburg sleep diary" OR "polysomnography" OR "polysomnography" OR "actigraphy" OR "actigraphy" ) AND ( "Generalized Anxiety Disorder" OR "Anxiety" OR "Anxiety Disorders" OR "Anxiety Disorders" OR "anxiety" OR "anxiety symptoms" OR "Social Anxiety" OR "panic disorder" OR "phobic disorders" OR "social anxiety disorder" OR "Social phobia" OR "specific phobia" OR "Agoraphobia" OR "Phobia related disorders" OR "Separation Anxiety Disorder" OR "Obsessive Compulsive Disorder" OR "Neurotic Disorders" OR "Screen for Anxiety Related Disorders" OR "Hamilton Anxiety Rating Scale" OR "Hamilton Anxiety Scale" OR "HAM A" OR "State Trait Anxiety Inventory" OR "STAI" OR "STAI S" OR "STAI 6" OR "Somatic Symptom Scale 8" OR "SSS 8" OR "Patient Health Questionnaire 15" OR "PHQ 15" OR "Generalized Anxiety Disorder 7 item" OR "7 item |  |
|  | Generalized Anxiety Disorder Scale" OR "GAD 7" OR "Hospital Anxiety and Depression Scale" OR "HADS" OR "Generalized Anxiety Disorder 2 item" OR "2 item Generalized Anxiety Disorder" OR "GAD 2") |  |

## Notes

### Competing Interest Statement

The authors have declared no competing interest.

